# Incidence and outcome of cancer in 16- to 17-year-old adolescents in a tertiary referral hospital

**DOI:** 10.1101/2021.05.31.21256303

**Authors:** Julia Ventelä, Atte Nikkilä, Arja Jukkola, Olli Lohi

## Abstract

**BACKGROUND:** Adolescents have a unique cancer profile that includes typical childhood cancers such as acute lymphoblastic leukemia (ALL) and adult-type cancers like melanoma and thyroid cancer. In Finland, adolescents above 16 years have been largely treated in adult oncology centers in contrast to many Western countries in which the minimum admittance age is 18 years. The aim of this study was to investigate characteristics, therapy and outcome of cancer in adolescents aged 16 to 17 years.

**MATERIALS AND METHODS:** This retrospective cohort study included adolescents aged 16 to under 18 years at the time of cancer diagnosis between 2000 and 2019 in Tampere University Hospital, a regional tertiary referral center. Clinical data were retrieved from hospital medical records and included diagnosis, clinical and laboratory parameters, therapy and outcome. In addition to standard descriptive statistical analyses, the Kaplan-Meier method and Cox regression modelling were applied to study the outcome and associated factors.

**RESULTS:** A total of 93 patients were diagnosed with a malignant tumor at the age of 16 or 17 years. Males were more often diagnosed with a cancer (62%), while non-CNS (non-central nervous system) solid cancers were the most common entities (76%) and Hodgkin lymphoma was the most common cancer diagnosis (31%). Seventy patients completed their therapy in the referral center and were followed up for a median of 5.4 years. The majority of patients were treated in the adult department (69%). Complete remission was achieved for 89% of patients, while 21% experienced tumor recurrence. The 5-year event-free survival was 65% (95% CI, 54% to 79%) and overall survival 82% (95% CI, 73% to 93%).

**CONCLUSIONS:** The majority of cancers in adolescents of 16 to 17 years of age are solid cancers. Patient outcome in our cohort was favorable and is in line with previously published data.

## Introduction

The term ‘child’ typically refers to a person under 18 years of age ^1^. Adolescence stretches between childhood and adulthood and it is often understood to comprise individuals from 10 to 24 years ^2^. Adolescents and young adults (AYA) refers to individuals aged between 15 and 39 years ^3^. Nowadays, the transition period from childhood to adulthood occupies a larger portion of life as earlier onset of puberty, delayed completion of education and later independence from parents is typical for young people in Western societies.^2^ This is relevant in the context of cancer care when considering the age at which a patient with cancer is treated in the pediatric or adult department.

Childhood cancers are classified according to histological and genetic type, whereas malignancies in adulthood are typically grouped by the primary location of the tumor. The International Classification of Childhood Cancers third edition (ICCC-3) is widely used to classify childhood cancers but is applicable for adolescents 15–19 years old ^4–5^. Overall, different solid cancers comprise approximately 60% of all cancers in this age group ^6^. The most common cancer types are hematopoietic cancers and lymphomas. Acute lymphoblastic leukemia (ALL) is the most common form of leukemia (86% of leukemias), while Hodgkin lymphoma is the most common lymphoma (75% of lymphomas) ^6,7^. In Finland, the incidence rate of cancer in children under 15 years of age is 17.8 per 100 000 individuals, while the respective number for adolescents of 15–19 years is 24.2 ^6^. The comparable incidence rates in adolescents are 20.5 and 23.5 in Europe and the US, respectively ^5,8^.

Survival rates of children and adolescents with cancer have improved significantly over time. In the 1960s, the majority of children with cancer died, while in the 2000s over 80% of patients can be cured ^9,10^. Survival rates differ significantly between children and adolescents. In 2000–2002, the proportion of survivors from lymphoid leukemia was 85% in children (0–14 years) and 50% in adolescents (15–24 years), and, for osteosarcoma, the comparable percentages were 77% and 60% ^11^. AYA patients with ALL have a worse outcome compared to children, but the prognosis has improved since implementation of pediatric-type protocols in the AYA group ^12,13^. The prognostic differences can be largely explained by the biological and genetic characteristics that are unique and specific to the AYA population ^14^.

In many European and Nordic countries, all patients under 18 years of age are treated in departments dedicated for children and/or adolescents. In contrast, only children below 16 years of age have been treated in pediatric units in Finland; therefore, 16- and 17-year-old patients have been predominantly managed in adult oncology or hematology departments. Access to multinational and pediatric-type protocols has been successful in treatment of AYA patients with ALL and clinical trial enrollment is generally associated with improvements in survival ^15,16^. Enrollment is lower among AYAs compared to the pediatric population. Many reasons for this are likely, such as the higher variability in cancer types and treatments, and lack of clinical trial availability for this age group. ^17–19^

In order to better understand the diagnostic and prognostic landscape of cancers among late teens, we scrutinized hospital medical records of patients with cancer who were 16 or 17 years old at the time of diagnosis in a Finnish tertiary referral hospital.

## Material and Methods

### Patient cohort

This retrospective cohort study was performed at Tampere University Hospital (TAUH) which serves as a tertiary referral center for patients in five counties in western and central Finland: Pirkanmaa, Kanta-Häme, Southern Ostrobothnia, Vaasa (up to 2012) and Päijät-Häme (up to 2017). The study population consisted of patients who were 16.0 to 17.99 years at the time of primary cancer diagnosis and diagnosed with a malignant tumor between 2000 and 2019 in TAUH. In total, our study included 93 patients with cancer, of which 70 completed therapy in the referral center and formed the main cohort (Figure 1 and Table S2). The study was approved by the research director of Pirkanmaa Hospital District in accordance with national research regulations.

**Figure 1.**
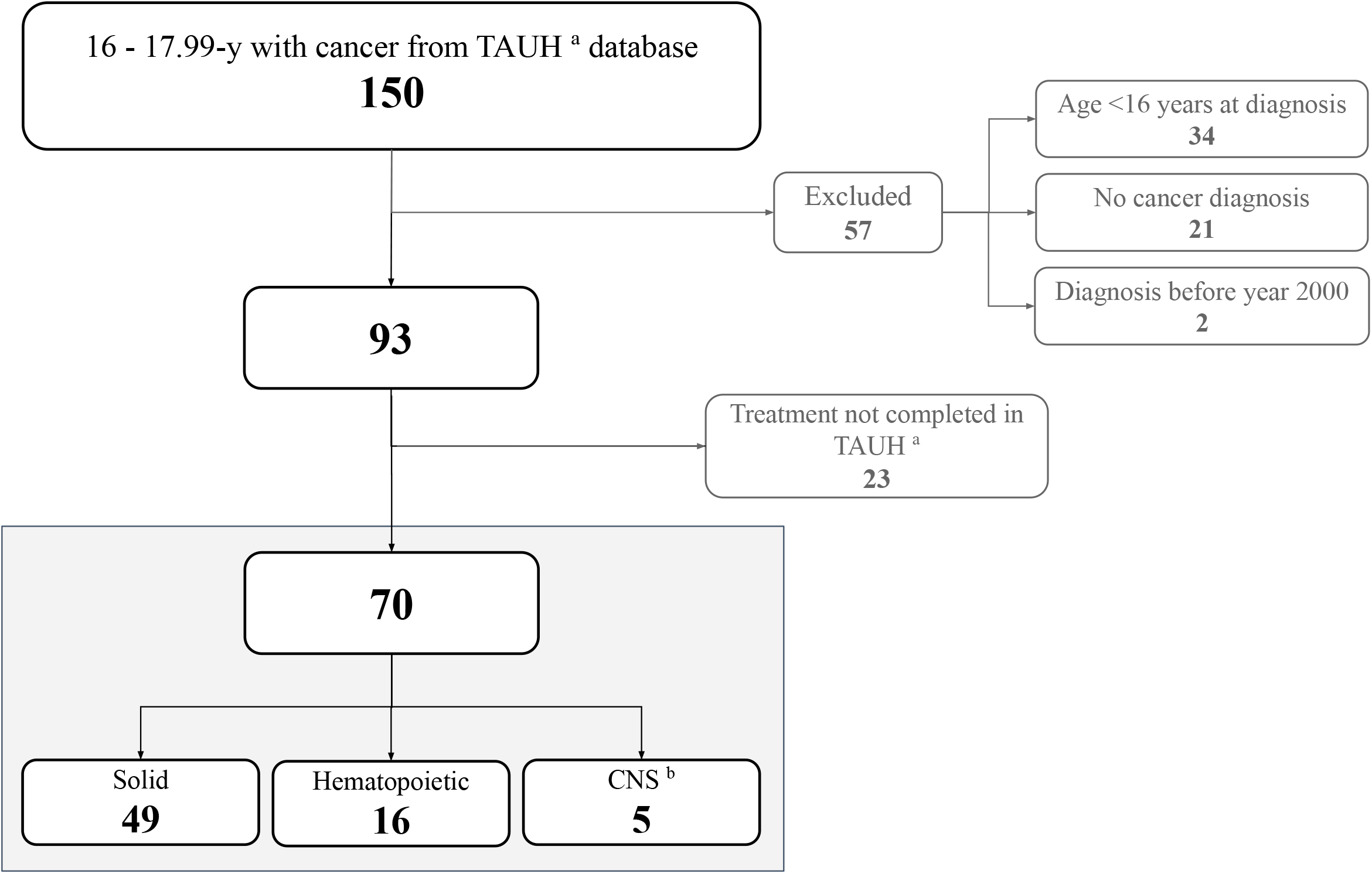
Flowchart of study subjects ^*a*^ *Tampere University Hospital* ^*b*^ *Central nervous system tumors*

### Data collection

Patient data were retrieved from electronic medical records (EMR) of TAUH. The screening criteria were the following: 1) The cancer was diagnosed between 1 January 2000 and 31 December 2019; 2) The ICD-10 codes were C00 to C97; and 3) The age of patient at the time of diagnosis was 16.0 to 17.99 years (Table 1). The patients were identified with a database query and clinical data was collected manually by a single researcher from the diagnosis until death or 1 June 2020. The data consisted of the precise diagnosis, the date of diagnosis, administered therapy and treatment protocol, the best response to treatment, survival status and the date of last follow-up. The best response was classified in four categories: complete remission, partial remission, stable disease or refractory/resistant disease. The definition for complete remission was the lack of signs of cancer in post-treatment imaging or sampling for solid cancers, and <5% of blasts in bone marrow for leukemias.

**Table 1.**
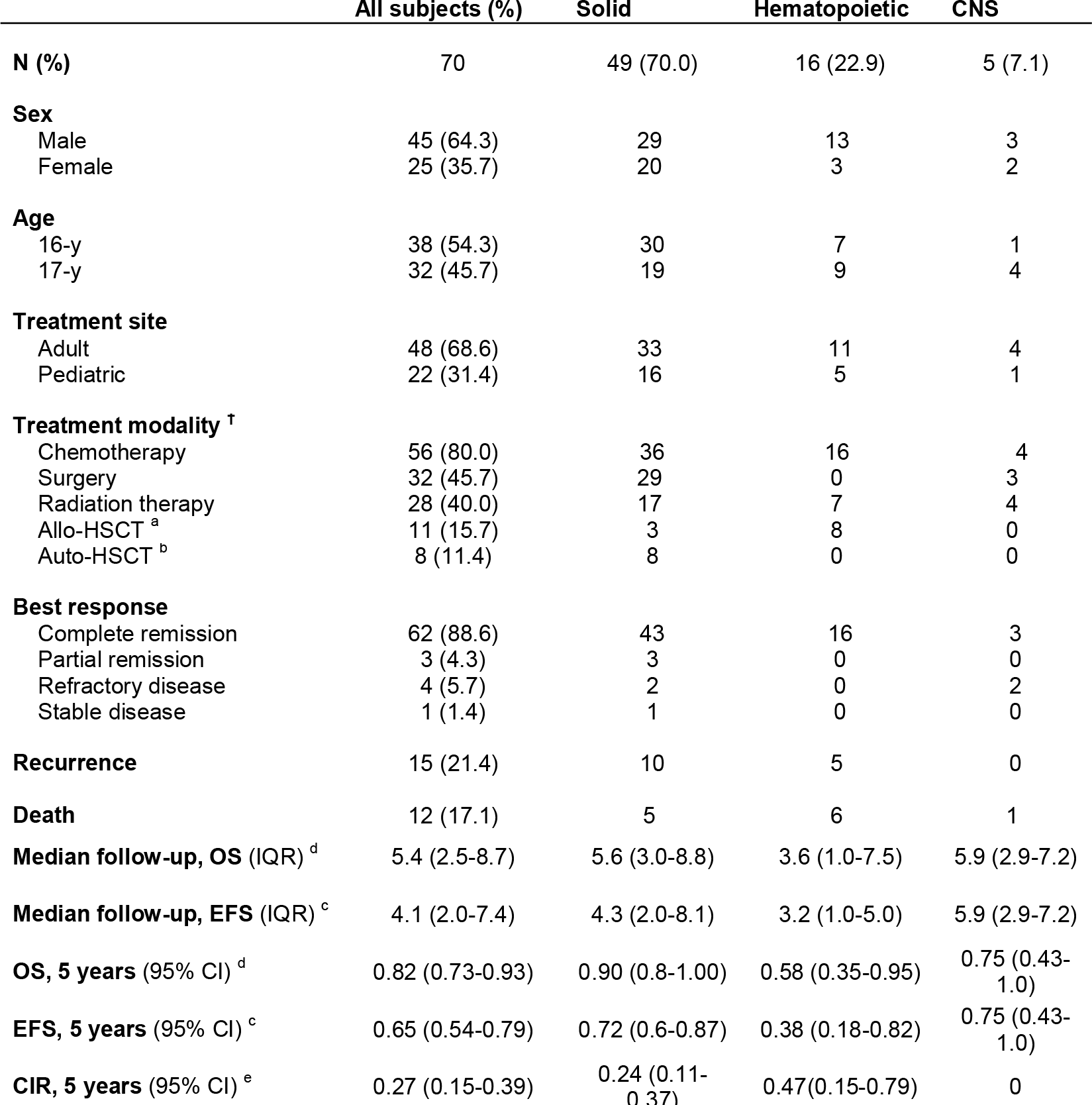
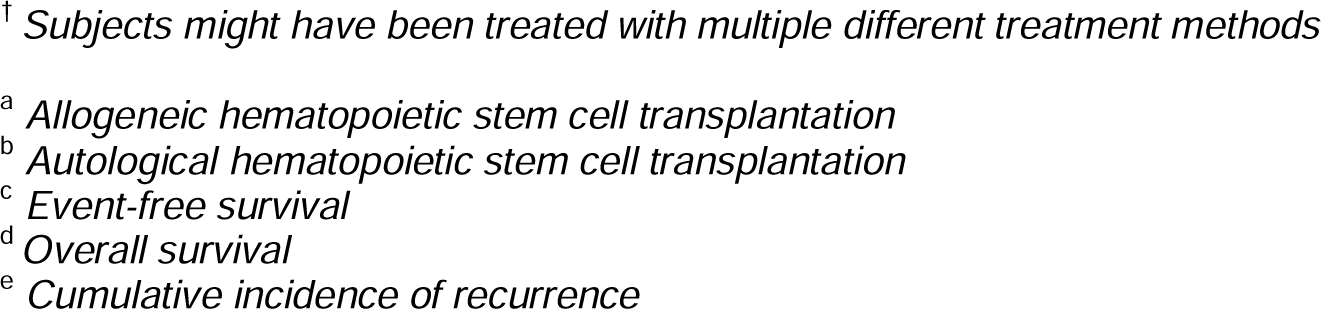
Characteristics of patients in the main cohort.

|  | All subjects (%) | Solid | Hematopoietic | CNS |
| --- | --- | --- | --- | --- |
| **N (%)** | 70 | 49 (70.0) | 16 (22.9) | 5 (7.1) |
| **Sex** |  |  |  |  |
| Male | 45 (64.3) | 29 | 13 | 3 |
| Female | 25 (35.7) | 20 | 3 | 2 |
| **Age** |  |  |  |  |
| 16-y | 38 (54.3) | 30 | 7 | 1 |
| 17-y | 32 (45.7) | 19 | 9 | 4 |
| **Treatment site** |  |  |  |  |
| Adult | 48 (68.6) | 33 | 11 | 4 |
| Pediatric | 22 (31.4) | 16 | 5 | 1 |
| **Treatment modality <sup>†</sup>** |  |  |  |  |
| Chemotherapy | 56 (80.0) | 36 | 16 | 4 |
| Surgery | 32 (45.7) | 29 | 0 | 3 |
| Radiation therapy | 28 (40.0) | 17 | 7 | 4 |
| Allo-HSCT <sup>a</sup> | 11 (15.7) | 3 | 8 | 0 |
| Auto-HSCT <sup>b</sup> | 8 (11.4) | 8 | 0 | 0 |
| **Best response** |  |  |  |  |
| Complete remission | 62 (88.6) | 43 | 16 | 3 |
| Partial remission | 3 (4.3) | 3 | 0 | 0 |
| Refractory disease | 4 (5.7) | 2 | 0 | 2 |
| Stable disease | 1 (1.4) | 1 | 0 | 0 |
| **Recurrence** | 15 (21.4) | 10 | 5 | 0 |
| **Death** | 12 (17.1) | 5 | 6 | 1 |
| **Median follow-up, OS (IQR) <sup>d</sup>** | 5.4 (2.5-8.7) | 5.6 (3.0-8.8) | 3.6 (1.0-7.5) | 5.9 (2.9-7.2) |
| **Median follow-up, EFS (IQR) <sup>c</sup>** | 4.1 (2.0-7.4) | 4.3 (2.0-8.1) | 3.2 (1.0-5.0) | 5.9 (2.9-7.2) |
| **OS, 5 years (95% CI) <sup>d</sup>** | 0.82 (0.73-0.93) | 0.90 (0.8-1.00) | 0.58 (0.35-0.95) | 0.75 (0.43-1.0) |
| **EFS, 5 years (95% CI) <sup>c</sup>** | 0.65 (0.54-0.79) | 0.72 (0.6-0.87) | 0.38 (0.18-0.82) | 0.75 (0.43-1.0) |
| **CIR, 5 years (95% CI) <sup>e</sup>** | 0.27 (0.15-0.39) | 0.24 (0.11-0.37) | 0.47(0.15-0.79) | 0 |

<sup>†</sup> *Subjects might have been treated with multiple different treatment methods*
<sup>a</sup> *Allogeneic hematopoietic stem cell transplantation*
<sup>b</sup> *Autological hematopoietic stem cell transplantation*
<sup>c</sup> *Event-free survival*
<sup>d</sup> *Overall survival*
<sup>e</sup> *Cumulative incidence of recurrence*

For the characteristic variables, the cancers were grouped into diagnostic categories according to type of tumor, i.e. solid, central nervous system (CNS) or hematopoietic malignancy.

### Data analysis and statistics

R software version 3.6.3 was used for statistical analyses. All calculated p-values are two-sided with 95% confidence intervals and p-values <0.05 were considered statistically significant. Overall survival (OS) was defined as the time from the diagnosis to death or to the last follow-up date, and event-free survival (EFS) was defined as the time from diagnosis to the first event (relapse, death or second malignancy) or to the last follow-up date. In our data, no second malignancies were registered. OS and EFS were estimated using the Kaplan-Meier method and the Log-rank test was used to evaluate the significance of difference between different groups. A Cox proportional hazard regression model was applied to estimate hazard ratios for outcome of the following covariates: sex, age, cancer type and treatment ward. We estimated cumulative incidence of relapse and disease recurrence by using the Kaplan-Meier method and Gray’s test with death as a competing event ^20^.

## Results

### Characteristics of the patients

A total of 93 patients aged 16 to 17 years were diagnosed with a malignant tumor between 2000 and 2019 in TAUH and clinics in its catchment area. Annually, a mean of 4.6 cases were diagnosed with a majority being males (62%) (Figure S1 and Table S1). Tumors were classified according to the ICCC-3 classification and the most common were Hodgkin lymphoma (29 cases; 31%), ALL (13 cases; 14%) and gonadal germ cell tumors (9 cases; 10%) (Table S1). While ALL was the most frequent hematopoietic malignancy, CNS neoplasms (5 cases; 5%) included diagnoses such as astrocytoma, ganglioglioma, medulloblastoma and intracranial germ cell tumors.

Out of 93 patients, 70 completed their treatment in TAUH and comprised the main cohort of the study (Table 1 and Table 2). The age spread was approximately even (54.3% 16 years; 45.7% 17 years). Of the 49 patients with solid cancers, 34.7% had stage one, 22.4% stage two, 12.2% stage three and 30.6% stage four disease. None of the leukemia patients had involvement of the central nervous system at diagnosis.

**Table 2.** Classification of patients according to the International Childhood Cancer Classification category (ICCC-3).

| ICCC-3 category | N (%) |
| --- | --- |
| **All subjects** | 70 |
| **I. Leukemia** | 16 (23) |
| Ia. ALL | 13 |
| Ib. AML | 3 |
| **II. Lymphoma** | 20 (29) |
| IIa. Hodgkin Lymphoma | 18 |
| IIb. Non-Hodgkin Lymphoma (non-Burkitt) | 1 |
| IIc. Burkitt Lymphoma | 1 |
| **III. CNS tumor <sup>a</sup>** | 3 (4) |
| IIIb. Astrocytoma | 1 |
| IIIc. Embryonal tumor | 1 |
| IIId. Glioma | 1 |
| **IV. Neuroblastoma** | 0 (0) |
| **V. Retinoblastoma** | 0 (0) |
| **VI. Renal tumor** | 1 (1) |
| VIa. Nephroblastoma | 1 |
| **VII. Hepatic tumor** | 0 (0) |
| **VIII. Malignant bone tumor** | 8 (11) |
| VIIIa. Osteosarcoma | 4 |
| VIIIb. Chondrosarcoma | 1 |
| VIIIc. Ewing sarcoma | 3 |
| **IX. Soft tissue and extraosseous sarcoma** | 5 (7) |
| IXa. Rhabdomyosarcoma | 1 |
| IXb. Fibrosarcoma | 1 |
| IXd. Other soft tissue sarcoma | 3 |
| **X. Germ cell tumor** | 8 (11) |
| Xa. Intracranial germ cell tumor | 2 |
| Xc. Gonadal germ cell tumor | 6 |
| **XI. Other malign. epithelial tumors and melanoma** | 9 (13) |
| XIb. Thyroid carcinoma | 4 |
| XIe. Skin carcinoma | 2 |
| XIf. Other and unspecified | 3 |

<sup>a</sup> Central nervous system

### Treatment and evaluation of response

The majority of the patients were treated at the adult oncology or hematology department (48 cases; 68.6 %) (Table 1). Malignancies treated at the pediatric department consisted mainly of ALL (5), Hodgkin lymphomas (5), and bone tumors (two osteosarcomas and three Ewing sarcomas).

Chemotherapy was the most frequent treatment modality (56 cases; 80.0%), followed by surgery (32 cases; 45.7%) and radiation therapy (28 cases; 40.0%). Allogeneic hematopoietic stem cell transplantation (HSCT) was performed for 11 cases (15.7%), whereas autologous hematopoietic stem cell support was applied in eight cases (11.4%).

The majority of the patients achieved complete remission (CR) (62 cases; 88.6%) (Table 1). Out of 22 patients who were treated in the pediatric department, 20 (91%) patients achieved CR, whereas 42 of the 48 patients (88%) treated at the adult department met the criteria of CR. Altogether, 15 patients (21.4%) experienced at least one relapse after achieving CR, of which ten were recurrences of solid cancers and five relapses of hematopoietic malignancies. None of the five patients with a CNS tumor experienced a recurrence.

### Survival of patients and frequency of recurrence

For all cancers, the median follow-up time was 5.4 years (IQR 2.5-8.7) and estimated EFS at 5-years was 65% (95% CI, 54% to 79%) (Figure S2A and Table 1) and OS 82% (95% CI, 73% to 93%) (Figure 2A and Table 1). The OS for males was worse than for females (73% vs 100%, p=0.041) (Figure 2B). The type of cancer associated with survival (p=0.015) (Figure 2C): hematopoietic malignancies had a 5-year OS of 58% (95% CI, 35% to 95%), while patients with solid cancers had an OS of 90.3% (95% CI, 82% to 100%) (Table 1). When looking at individual cancer types, the ALL patients had a 5-year OS of 61% (95% CI, 38% to 100%), and Hodgkin lymphoma patients had a 5-year OS of 92% (95% CI, 79% to 100%). In EFS analysis, we found no statistically significant differences between sexes (p=0.50) or different tumor types (p=0.21) (Figure S2B and S2C).

**Figure 2.**
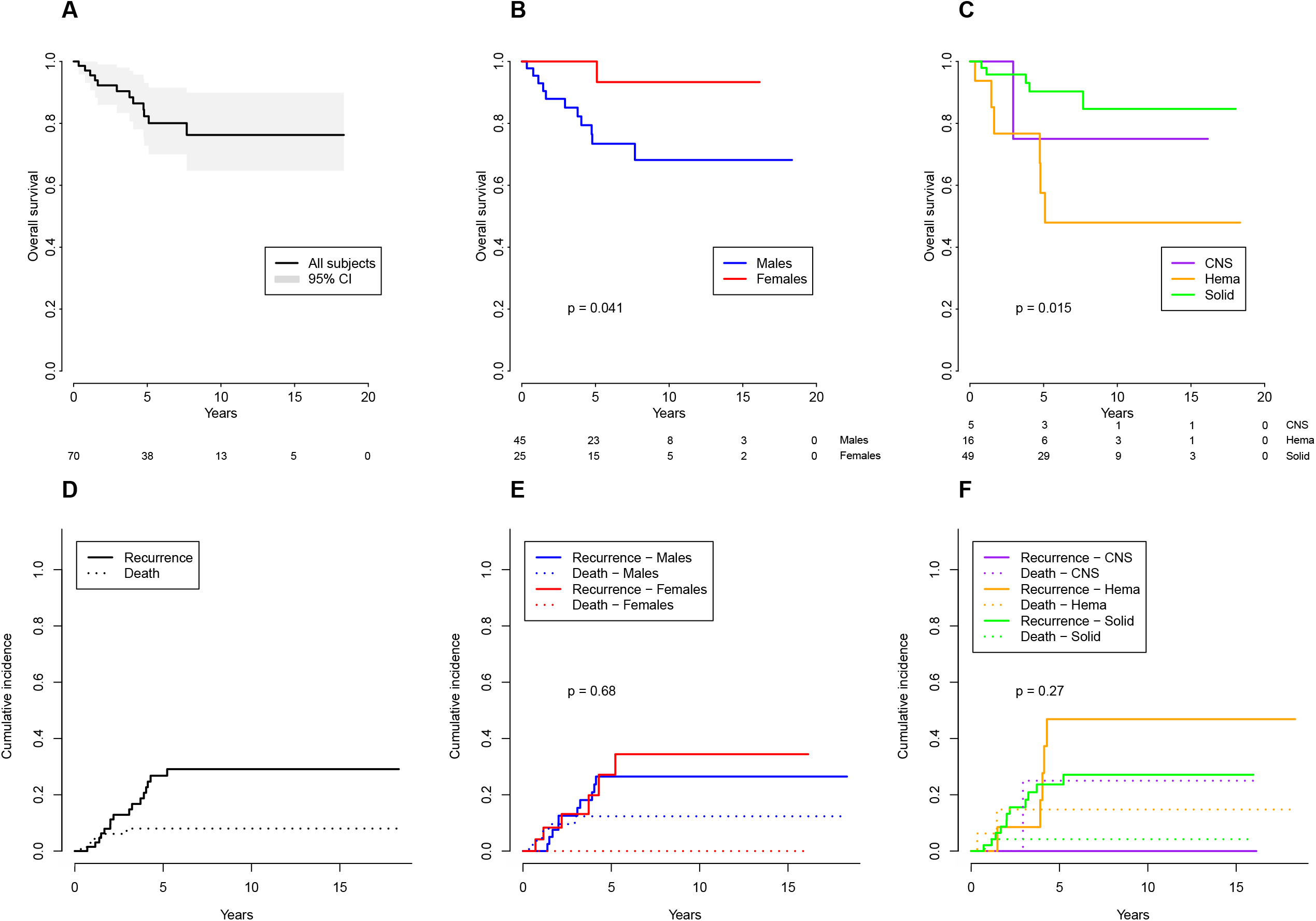
Overall survival for all patients (A), and stratified by either sex (B) or cancer type (C) and cumulative incidence of recurrence and death for all patients (D), and stratified by either sex (E) or cancer type (F). *The shaded area in the panel A represents the 95% confidence intervals. The p-values were calculated using the logrank test. The p-values in the panels E (p=0*.*68) and F (p=0*.*27) are calculated for the cumulative incidence of recurrence*.

We next investigated the impact of covariates on survival by using Cox regression modelling. In univariate and multivariate models, patients with hematopoietic malignancies had the worst outcome (Table 3). No significantly increased hazard for death was observed for sex, age, cancer type, or the treatment site.

**Table 3.** Cox proportional hazards model for overall survival.

|  | N | Multivariate |  |  |  | Univariate |  |  |
| --- | --- | --- | --- | --- | --- | --- | --- | --- |
|  |  | HR <sup>a</sup> | 95% CI <sup>b</sup> | p |  | HR <sup>a</sup> | 95% CI <sup>b</sup> | p |
| **All subjects** | 70 |  |  |  |  |  |  |  |
| **Sex** |  |  |  |  |  |  |  |  |
| Male | 45 | 1.00<sup>†</sup> |  |  |  | 1.00<sup>†</sup> |  |  |
| Female | 25 | 0.21 | 0.03-1.69 | 0.14 |  | 0.16 | 0.02-1.21 | 0.08 |
| **Age** |  |  |  |  |  |  |  |  |
| 16-y | 38 | 1.00<sup>†</sup> |  |  |  | 1.00<sup>†</sup> |  |  |
| 17-y | 32 | 0.99 | 0.23-4.2 | 0.99 |  | 1.9 | 0.61-6.10 | 0.26 |
| **Tumor type** |  |  |  |  |  |  |  |  |
| Solid | 49 | 1.00<sup>†</sup> |  |  |  | 1.00<sup>†</sup> |  |  |
| Hematopoietic | 16 | 4.44 | 1.22-16.2 | 0.024* |  | 4.89 | 1.49-16.1 | 0.009* |
| CNS | 5 | 2.23 | 0.23-21.2 | 0.49 |  | 2.01 | 0.23-17.2 | 0.52 |
| **Treatment site** |  |  |  |  |  |  |  |  |
| Adult | 48 | 1.00<sup>†</sup> |  |  |  | 1.00<sup>†</sup> |  |  |
| Pediatric | 22 | 0.49 | 0.10-2.5 | 0.30 |  | 0.64 | 0.17-2.37 | 0.50 |

The Cox analyses were calculated with R (v. 3.6.3) with function `coxph()` from library `survival`. The proportional hazards (PH) assumption was assessed by the `cox.zph()` function.
<sup>†</sup> The reference category.
\*Statistically significant p-value, <0.05
<sup>a</sup> HR=hazard ratio
<sup>b</sup> CI = confidence interval

Cumulative incidence of recurrence (CIR) was calculated with the Gray’s test by using death before recurrence as a competing factor. Among all patients, CIR at 5 years was 27% (95% CI, 15% to 39%) (Table 1). The CIR was the highest among hematopoietic malignancies (p=0.27), and did not differ between sexes (p=0.68) (Figure 2E and 2F).

## Discussion

Tumor biology is characteristic for each tumor and varies across diagnoses and age groups. Therapy response depends primarily on tumor biology and dissemination, but is also affected by the type and intensity of the administered therapy. In Finland, traditionally adolescents of 16 and 17 years of age have been largely managed in adult departments. We investigated this specific subgroup of patients in a retrospective setting to understand the diagnostic and prognostic characteristics of these patients in a tertiary referral hospital that serves a population of over one million people. The spectrum of diagnoses mirrored the transitional phase with a diminishing number of hematopoietic malignancies and higher proportion of solid cancers. Overall, the patient outcome was favorable irrespective of treatment site.

Hodgkin lymphoma, ALL, soft tissue sarcoma and bone tumors were the most common diagnoses in our patient cohort. Typical young adult-type cancers, such as thyroid cancer, melanoma and testicular germ cell tumors were seen as well. The cancer types in our data align well with the literature in terms of the cancer types typical within the AYA population ^21,22^.

Over three quarters of patients were long-term survivors with 5-year OS standing at 82%. Outcome varied depending on the type of cancer and malignancies of hematopoietic origin had the poorest prognosis. On the other hand, over 90% of Hodgkin lymphoma patients were long-term survivors. The survival rates in our cohort closely resemble the outcome data from analysis of childhood cancer patients in Europe and Finland ^9,11^. At the European level, adolescents aged 15 to 24 years with Hodgkin lymphoma had a 5-year OS of 93% and with ALL 49.5% ^23^.

Males with cancer had an unfavorable outcome in our cohort. This is largely explained by the biased gender distribution among hematopoietic cancer patients, where 81% of patients were males. Adolescent patients with hematopoietic malignancies had the poorest 5-year OS, whereas the patients with solid cancers had the most favourable prognosis. No differences were seen between the sexes in EFS, indicating that females were more likely to survive from recurrences than males.

Out of 70 cases that were treated in the referral center, 31% were managed in the pediatric department. Cases treated there included, e.g., leukemias, lymphomas, bone tumors, and testicular cancers. We did not see any significant difference in the prognosis of patients in terms of the treatment site as 91% and 88% of the patients who were treated in the pediatric or adult ward, respectively, achieved CR. In the Cox regression model, a non-significant trend suggestive of a better outcome in patients treated at the pediatric ward was observed.

Our study has several strengths: the patient data is population-based, collected from a single tertiary referral center, and all the diagnoses were confirmed by histopathology. One limitation is the relatively low number of patients. Likewise, although the recruitment period was long and lasted up to 20 years, the median follow-up of the patients was much shorter and reflects the transitional phase of life of young patients. In the future, we plan to expand the study population in terms of age groups and geographical spread to obtain a broader view of adolescent malignancies, their treatment and outcome.

## Data Availability

The authors are unable to share the raw data due to the European Union General Data Protection Regulation (GDPR) and Finland's specific legislation on sharing of protected personal data.

## Data availability

The authors are unable to share the raw data due to the European Union General Data Protection Regulation (GDPR) and Finland’s specific legislation on sharing of protected personal data ^24,25^.

## Supplemental material

**Figure S1**. Histogram of annual incidence of adolescent cancer diagnoses in Tampere University Hospital (2000-2019).

**Figure S2**. Event free survival for all patients (A), and stratified by either sex (B) or cancer type (C).

**Table S1**. Classification according to ICCC-3 for adolescent cancer patients who were diagnosed in Tampere University Hospital.

**Table S2**. Excluded subjects stratified by sex, exclusion criteria and cancer type.

## Notes

**Disclosure of interest**. The authors report no conflicting interests.

### Competing Interest Statement

The authors have declared no competing interest.

### Funding Statement

No external funding was received.

### Author Declarations

The study protocol was approved by Tampere University Hospital chief physician.

