## Supplementary material for "Incidence and outcome of cancer in 16- to 17-year-old adolescents in a tertiary referral hospital": Table S1

**Table S1.**  Sex, age and ICCC3 classification for patients aged 16–17 at diagnosis in TAUH ^a^ and its catchment area.

|  | **N (%)** |
| --- | --- |
| **All subjects** | 93 |
| **Sex** |  |
| Female | 35 (37.6) |
| Male | 58 (62.3) |
| **Age** |  |
| 16-y | 47 (50.5) |
| 17-y | 46 (49.4) |
| **I. Leukemia** | 17 (18.2) |
| Ia. ALL | 13 |
| Ib. AML | 4 |
| **II. Lymphoma** | 34 (36.6) |
| IIa. Hodgkin Lymphoma | 29 |
| IIb. Non-Hodgkin Lymphoma (non-Burkitt) | 4 |
| IIc. Burkitt Lymphoma | 1 |
| **III. CNS** **tumor** ^b^ | 3 (3.2) |
| IIIb. Astrocytoma | 1 |
| IIIc. Embryonal tumor | 1 |
| IIId. Glioma | 1 |
| **IV. Neuroblastoma**  **V. Retinoblastoma**  **VI. Renal tumor** | 0 (0)  0 (0)  1 |
| VIa. Nephroblastoma | 1 |
| **VII. Hepatic tumor** | 0 (0) |
| **VIII. Malignant bone tumor** | 10 (10.8) |
| VIIIa. Osteosarcoma | 5 |
| VIIIb. Chondrosarcoma | 1 |
| VIIIc. Ewing sarcoma | 4 |
| **IX. Soft tissue and extraosseous sarcoma** | 6 (6.5) |
| IXa. Rhabdomyosarcoma | 1 |
| IXb. Fibrosarcoma | 1 |
| IXd. Other soft tissue sarcoma | 4 |
| **X. Germ cell tumor** | 11 (11.8) |
| Xa. Intracranial germ cell tumor | 2 |
| Xc. Gonadal germ cell tumor | 9 |
| **XI. Other malig. epithelial tumors and melanomas** | 11 (11.8) |
| XIb. Thyroid carcinoma | 4 |
| XId. Melanoma | 1 |
| XIe. Skin carcinoma | 2 |
| XIf. Other and unspecified | 4 |

*^a^ Tampere University Hospital*

*^b^ Central nervous system*
