## Supplementary material for "Incidence and outcome of cancer in 16- to 17-year-old adolescents in a tertiary referral hospital": Table S2

**Table S2.** Subjects that were excluded from the analysis stratified by sex, exclusion criteria and tumor class.

|  | **All subjects (%)** | **Female** | **Male** |
| --- | --- | --- | --- |
| **N (%)** | 80 | 39 (48.8) | 41 (51.3) |
| **Exclusion criteria** |  |  |  |
| Age under 16 years | 34 (42.5) | 18 | 16 |
| Treatments not in TAUH ^a^ | 23 (28.8) | 10 | 13 |
| Final diagnosis other than cancer ^†^ | 21 (26.3) | 10 | 11 |
| Diagnosis before year 2000 | 2 (2.5) | 1 | 1 |
| **Tumor classification** ^‡^ |  |  |  |
| Solid tumors | 47 (79.7) | 25 | 22 |
| Hematopoietic | 7 (11.9) | 3 | 4 |
| CNS tumors | 5 (8.5) | 1 | 4 |

^†^ *The diagnosis became more accurate later in the treatments or was incorrectly reported in the first place.*

^‡^ *The number of subjects with no cancer diagnosis was subtracted from the denominator of the percentages.*

^a^ *Tampere University Hospital*
