## Supplementary figures and images for "Incidence and outcome of cancer in 16- to 17-year-old adolescents in a tertiary referral hospital"

### Figure S1

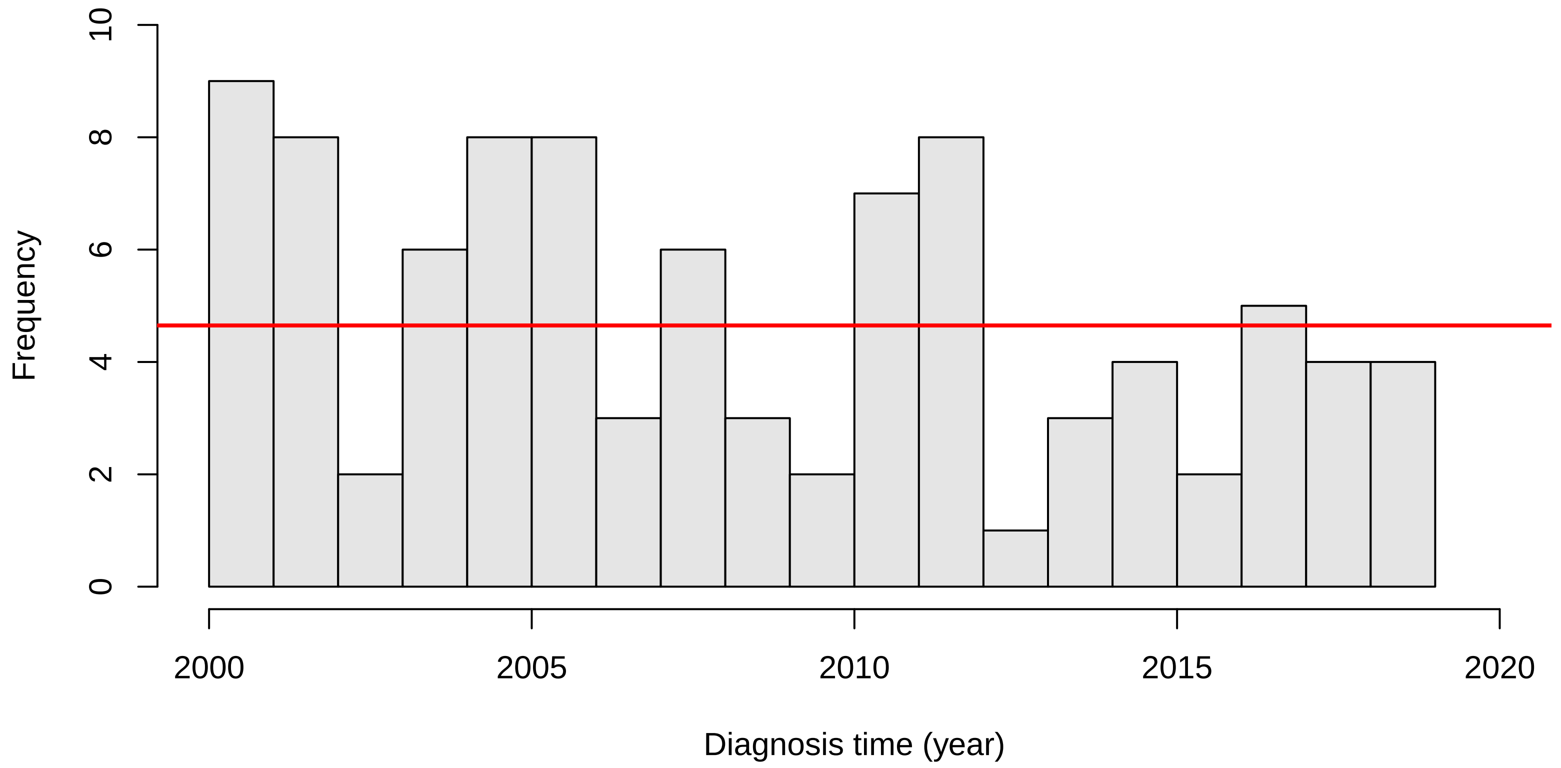

### Figure S1

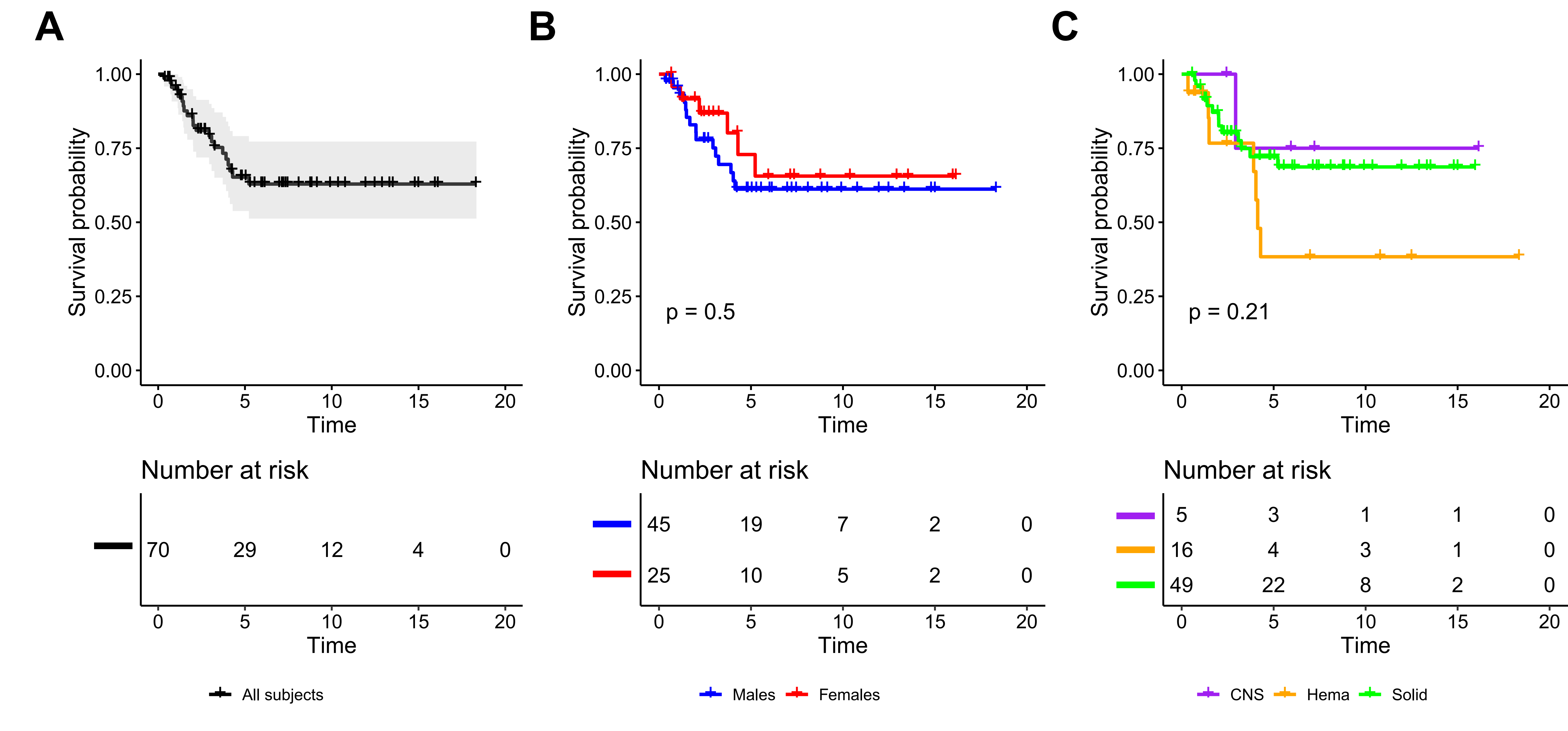
